# A deep learning algorithm using CT images to screen for Corona Virus Disease (COVID-19)

**DOI:** 10.1101/2020.02.14.20023028

**Authors:** Shuai Wang, Bo Kang, Jinlu Ma, Xianjun Zeng, Mingming Xiao, Jia Guo, Mengjiao Cai, Jingyi Yang, Yaodong Li, Xiangfei Meng, Bo Xu

**Affiliations:** Department of Molecular Radiation Oncology, National Clinical Research Center for Cancer, Key Laboratory of Cancer Prevention and Therapy, Key Laboratory of Breast Cancer Prevention and Therapy, Ministry of Education, Tianjin Clinical Research Center for Cancer, Tianjin Medical University Cancer Institute and Hospital, Tianjin 300060, China; College of Intelligence and Computing, Tianjin University, Tianjin 300350, China; National Supercomputer Center in Tianjin, Tianjin 300457, China; Department of Radiation Oncology, First Affiliated Hospital, Xi’an Jiaotong University, Xi’an, China; Department of Radiology, Nanchang University First Hospital, Nanchang, China; Department of Radiology, No.8 Hospital, Xi’an Medical College, Xi’an, China

**Keywords:** COVID-19, Computed Tomography, Artificial Intelligence, Deep Learning, Diagnosis

## Abstract

**Background:** The outbreak of Severe Acute Respiratory Syndrome Coronavirus 2 (SARS-COV-2) has caused more than 2.5 million cases of Corona Virus Disease (COVID-19) in the world so far, with that number continuing to grow. To control the spread of the disease, screening large numbers of suspected cases for appropriate quarantine and treatment is a priority. Pathogenic laboratory testing is the gold standard but is time-consuming with significant false negative results. Therefore, alternative diagnostic methods are urgently needed to combat the disease. Based on COVID-19 radiographical changes in CT images, we hypothesized that Artificial Intelligence’s deep learning methods might be able to extract COVID-19’s specific graphical features and provide a clinical diagnosis ahead of the pathogenic test, thus saving critical time for disease control.

**Methods and Findings:** We collected 1,065 CT images of pathogen-confirmed COVID-19 cases (325 images) along with those previously diagnosed with typical viral pneumonia (740 images). We modified the Inception transfer-learning model to establish the algorithm, followed by internal and external validation. The internal validation achieved a total accuracy of 89.5% with specificity of 0.88 and sensitivity of 0.87. The external testing dataset showed a total accuracy of 79.3% with specificity of 0.83 and sensitivity of 0.67. In addition, in 54 COVID-19 images that first two nucleic acid test results were negative, 46 were predicted as COVID-19 positive by the algorithm, with the accuracy of 85.2%.

**Conclusion:** These results demonstrate the proof-of-principle for using artificial intelligence to extract radiological features for timely and accurate COVID-19 diagnosis.

**Author summary:** To control the spread of the COVID-19, screening large numbers of suspected cases for appropriate quarantine and treatment measures is a priority. Pathogenic laboratory testing is the gold standard but is time-consuming with significant false negative results. Therefore, alternative diagnostic methods are urgently needed to combat the disease. We hypothesized that Artificial Intelligence’s deep learning methods might be able to extract COVID-19’s specific graphical features and provide a clinical diagnosis ahead of the pathogenic test, thus saving critical time. We collected 1,065 CT images of pathogen-confirmed COVID-19 cases along with those previously diagnosed with typical viral pneumonia. We modified the Inception transfer-learning model to establish the algorithm. The internal validation achieved a total accuracy of 89.5% with specificity of 0.88 and sensitivity of 0.87. The external testing dataset showed a total accuracy of 79.3% with specificity of 0.83 and sensitivity of 0.67. In addition, in 54 COVID-19 images that first two nucleic acid test results were negative, 46 were predicted as COVID-19 positive by the algorithm, with the accuracy of 85.2%. Our study represents the first study to apply artificial intelligence to CT images for effectively screening for COVID-19.

## Introduction

The outbreak of atypical and person-to-person transmissible pneumonia caused by the severe acute respiratory syndrome coronavirus 2 (SARS-COV-2, also known as 2019-nCov) has caused a global alarm. There have been more than 2.5 million confirmed cases of the Corona Virus Disease (COVID-19) in the world, as of April 23, 2020. About 16-21% of people with the virus in China have become severely ill with a 2-3% mortality rate. With the most recent estimated viral reproduction number (R0), the average number of other people that an infected individual will transmit the virus to in a completely non-immune population, stands at about 3.77 [1], indicating that a rapid spread of the disease is imminent. It is crucial to identify infected individuals as early as possible for quarantine and treatment procedures.

The diagnosis of COVID-19 relies on the following criteria: clinical symptoms, epidemiological history and positive CT images, as well as positive pathogenic testing. The clinical characteristics of COVID-19 include respiratory symptoms, fever, cough, dyspna, and pneumonia [3-6]. However, these symptoms are nonspecific, as there are isolated cases where, for example, in an asymptomatic infected family a chest CT scan revealed pneumonia and the pathogenic test for the virus came back positive. Once someone is identified as a PUI (person under investigation), lower respiratory specimens, such as bronchoalveolar lavage, tracheal aspirate or sputum, will be collected for pathogenic testing. This laboratory technology is based on real-time RT-PCR and sequencing of nucleic acid from the virus [7,8]. Since the beginning of the outbreak, the efficiency of nucleic acid testing has been dependent on several rate-limiting factors, including availability and quantity of the testing kits in the affected area. More importantly, the quality, stability and reproducibility of the detection kits are questionable. The impact of methodology, disease stages, specimen collection methods, nucleic acid extraction methods, and the amplification system are all determinant factors for the accuracy of test results. Conservative estimates of the detection rate of nucleic acid are low (between 30-50%) [7,8,9], and tests need to be repeated several times in many cases before they can be confirmed.

Radiological imaging is also a major diagnostic tool for COVID-19. The majority of COVID-19 cases have similar features on CT images including ground-glass opacities in the early stage and pulmonary consolidation in the late stage. There is also sometimes a rounded morphology and a peripheral lung distribution [6,10]. Although typical CT images may help early screening of suspected cases, the images of various viral pneumonias are similar and they overlap with other infectious and inflammatory lung diseases. Therefore, it is difficult for radiologists to distinguish COVID-19 from other viral pneumonias. Artificial Intelligence involving medical imaging deep-learning systems has been developed in image feature extraction, including shape and spatial relation features. Specifically, Convolutional Neural Network (CNN) has been proven in feature extraction and learning. CNN was used to enhance low-light images from high-speed video endoscopy with the limited training data being just 55 videos [11]. Also, CNN has been applied to identify the nature of pulmonary nodules via CT images, the diagnosis of pediatric pneumonia via chest X-ray images, automated precising and labeling of polyps during colonoscopic videos, cystoscopic image recognition extraction from videos [12-15].

There are a number of features for identifying viral pathogens on the basis of imaging patterns, which are associated with their specific pathogenesis [16]. The hallmarks of COVID-19 are bilateral distribution of patchy shadows and ground glass opacity in early stages. As the disease progresses, multiple ground glass and infiltrates in both lungs will appear [3]. These features are quite similar to typical viral pneumonia with only slight differences, which are difficult to be distinguished by radiologists. Based on this, we believed that CNN might help us identify unique features that might be difficult for visual recognition.

Hence, the purpose of our study was to evaluate the diagnostic performance of a deep learning algorithm using CT images to screen for COVID-19 during the influenza season. To test this notion, we retrospectively enrolled 1,065 CT images of pathogen-confirmed COVID-19 cases along with previously diagnosed typical viral pneumonia. Our results reported below demonstrate the proof-of-principle using the deep learning method to extract radiological graphical features for COVID-19 diagnosis.

## Methods and Materials

### Retrospective collection of datasets

We retrospectively collected CT images from 259 patients, in which the cohort includes 180 cases of typical viral pneumonia and the other 79 cases from three hospitals with confirmed nucleic acid testing of SARS-COV-2. In addition, we enrolled additional 15 COVID cases, in which first two nucleic acid tests were negative at initial diagnoses. Hospitals providing the images were Xi’an Jiaotong University First Affiliated Hospital (Center 1), Nanchang University First Hospital (Center 2) and Xi’an No.8 Hospital of Xi’an Medical College (Center 3). All CT images were de-identified before sending for analysis. This study is in compliance with the Institutional Review Board of each participating institutes. Informed consent was exempted by the IRB because of the retrospective nature of this study.

### Delineation of ROIs

To establish a binary model for distinguishing COVID-19 and typical pneumonia, we drew the Region of Interest (ROI) as input images for the training cohort and validation cohorts. We sketched the ROI from CT images based on features of COVID-19, such as small patchy shadows and interstitial changes in the early stage, multiple ground glass and infiltrates in both lungs in the progression stage, and delineated the ROIs on the CT images of other typical viral pneumonia such as pseudocavity, enlarged lymphnodes and multifocal GGO as the control. The ROIs were divided into three cohorts: one training cohort (n=320 from Center 1), one internal validation cohort (n=455 from Center 1) and one external validation cohort (n=290 from Center 2 and 3). For a ROI, it is sized approximately from 395*223 to 636*533 pixels.

### Overview of the proposed architecture

Our systematic pipeline for the prediction architecture is depicted in **Fig 1**. The architecture consists of three main processes: 1) Pre-processing of input images; 2) Feature extraction of ROI images and training; and 3) Classification with fully connected network and prediction of multiple classifiers. We built a transfer learning neural network based on the Inception network. The entire neural network can be roughly divided into two parts: the first part used a pre-trained inception network to convert image data into one-dimensional feature vectors, and the second part used a fully connected network and the main role is for classification prediction. ROI images from each case were preprocessed and inputted into the model for training. The number of various types of pictures in the training set is equal, with a total number of 320. The remaining CT pictures of each case were used for internal validation. The model training was iterated 15,000 times with a step size of 0.01.

**Fig 1.**
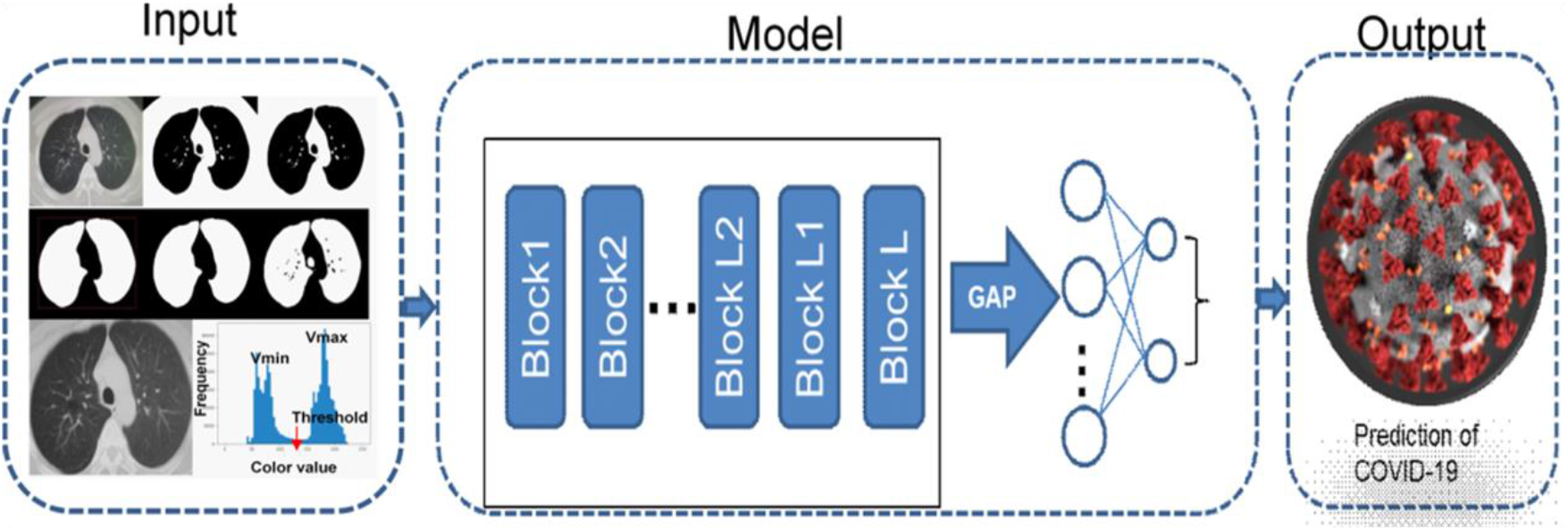
ROI images extraction and Deep Learning (DL) algorithm framework.

ROI images were extracted by the CV model and then trained using a modified Inception network to extract features. The full connection layer then makes a classification and prediction.

### Image preprocessing and feature extraction

Based on the signs of characteristic of pneumonia, ROI images were defined inflammatory lesions and extracted by our computer vision (CV) model following the steps. 1) Convert the image to grayscale. 2) Binarize grayscale. Because using the OSTU’s method directly may cause the threshold selection failure in the case of multi-peaks, the selection of the binarization threshold in this paper was based on the statistics of all pixel frequency histograms of the gray color values Vmin (80) and Vmax (200). The minimum frequency in the selection interval is threshold, and the interval of frequency statistics is five. 3) Background area filling. Using the flood filling method to expand the image by 1 black pixel, and fill the black pixels near the border with white. 4) Reverse color, find all the contour areas of the image, and keep the two largest contour areas as the two lung areas. 5) Take the smallest bounding rectangle of the lung area as the ROI frame and crop the original image to obtain the ROI images. The delineated ROIs were obtained for classification model building. We modified the typical Inception network, and fine-tuned the modified Inception (M-Inception) model with pre-trained weights. During the training phase, the original Inception part was not trained, and we only trained the modified part. The architecture of M-Inception is shown in **Table 1**. The difference between Inception and M-Inception in classification lies in the last fully-connected layers. We reduced the dimension of features before it was sent to the final classification layer. The training dataset made up of all those patches aforementioned. The Inception network is shown in **Table 1**.

**Table 1.**
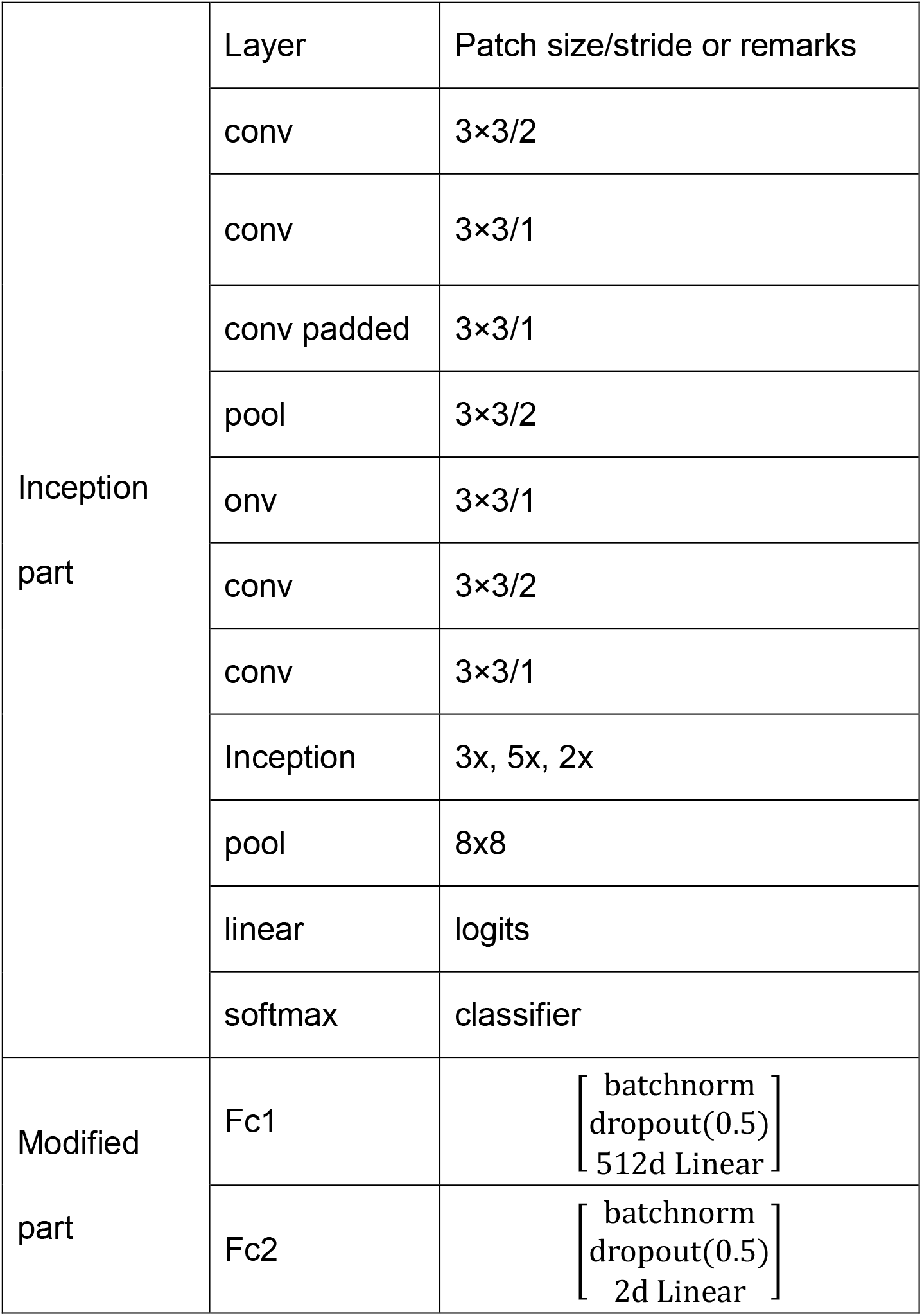
**The architecture of M-Inception**

|  |  |  |
| --- | --- | --- |
| Inception<br>part | Layer | Patch size/stride or remarks |
|  | conv | 3×3/2 |
|  | conv | 3×3/1 |
|  | conv padded | 3×3/1 |
|  | pool | 3×3/2 |
|  | conv | 3×3/1 |
|  | conv | 3×3/2 |
|  | conv | 3×3/1 |
|  | Inception | 3x, 5x, 2x |
|  | pool | 8x8 |
|  | linear | logits |
|  | softmax | classifier |
| Modified<br>part | Fc1 | $\begin{bmatrix} \text{batchnorm} \\ \text{dropout}(0.5) \\ 512\text{d Linear} \end{bmatrix}$ |
| | Fc2 | $\begin{bmatrix} \text{batchnorm} \\ \text{dropout}(0.5) \\ 2\text{d Linear} \end{bmatrix}$ |

### Prediction

After generating the features, the final step was to classify the pneumonia based on those features. Ensembling of classifiers was used to improve the classification accuracy. In this study, we adopted end-to-end learning to make the model convergence.

### Performance evaluation metrics

We compared the classification performance using Accuracy, Sensitivity, Specificity, Area Under Curve (AUC), Positive predictive value (PPV), Negative predictive value (NPV), F1 score and Youden Index. TP and TN represent the number of true positive or true negative samples. FP and FN mean the number of false positive or false negative samples. Sensitivity measures the ratio of positives that are correctly discriminated. Specificity measures the ratio of negatives that are correctly discriminated. AUC is an index to measure the performance of the classifier. NPV was used to evaluate the algorithm for screening, and PPV was the probability of getting a disease when the diagnostic index is positive. Youden Index was the determining exponent of the optimal bound. F1 score was a measure of the accuracy of a binary model. Additionally, performance was evaluated with F-measure (F1) to compare the similarity and diversity of performance. Kappa value measures the agreement between the CNN model prediction and the clinical report.

## Results

### Algorithm development

In order to develop a deep learning algorithm for the identification of viral pneumonia images, we initially enrolled 259 patients, in which the cohort includes 180 cases of typical viral pneumonia that were diagnosed previously before the COVID-19 outbreak. These patients are termed COVID-19 negative in the cohort. The other 79 cases were from the three hospitals with confirmed nucleic acid testing of SARS-COV-2, therefore termed COVID-19 positive. Two radiologists were asked to review the images and sketched 1,065 representative images (740 for COVID-19 negative and 325 for COVID-19 positive) for analysis (**Fig 2** is shown as an example). These images were randomly divided into a training set and a validation set. The model training was iterated for 15,000 times with a step size of 0.01. The training loss curve is shown in **Fig 3A**. 320 images (160 images from COVID-19 negative and 160 images from COVID-19 positive) were obtained to construct the model. To test the stability and generalization of the model, 455 images (COVID-19 ngegative 360 images and COVID-19 positive 95 images) were obtained for internal validation from Center 1 and 290 images (COVID-19 negative 220 images and COVID-19 positive 70 images) were obtained from Center 2 and 3 for external validation. The model training was also iterated for 15,000 times with a step size of 0.01. The training loss curve is shown in **Fig 3B**.

**Fig 2.**
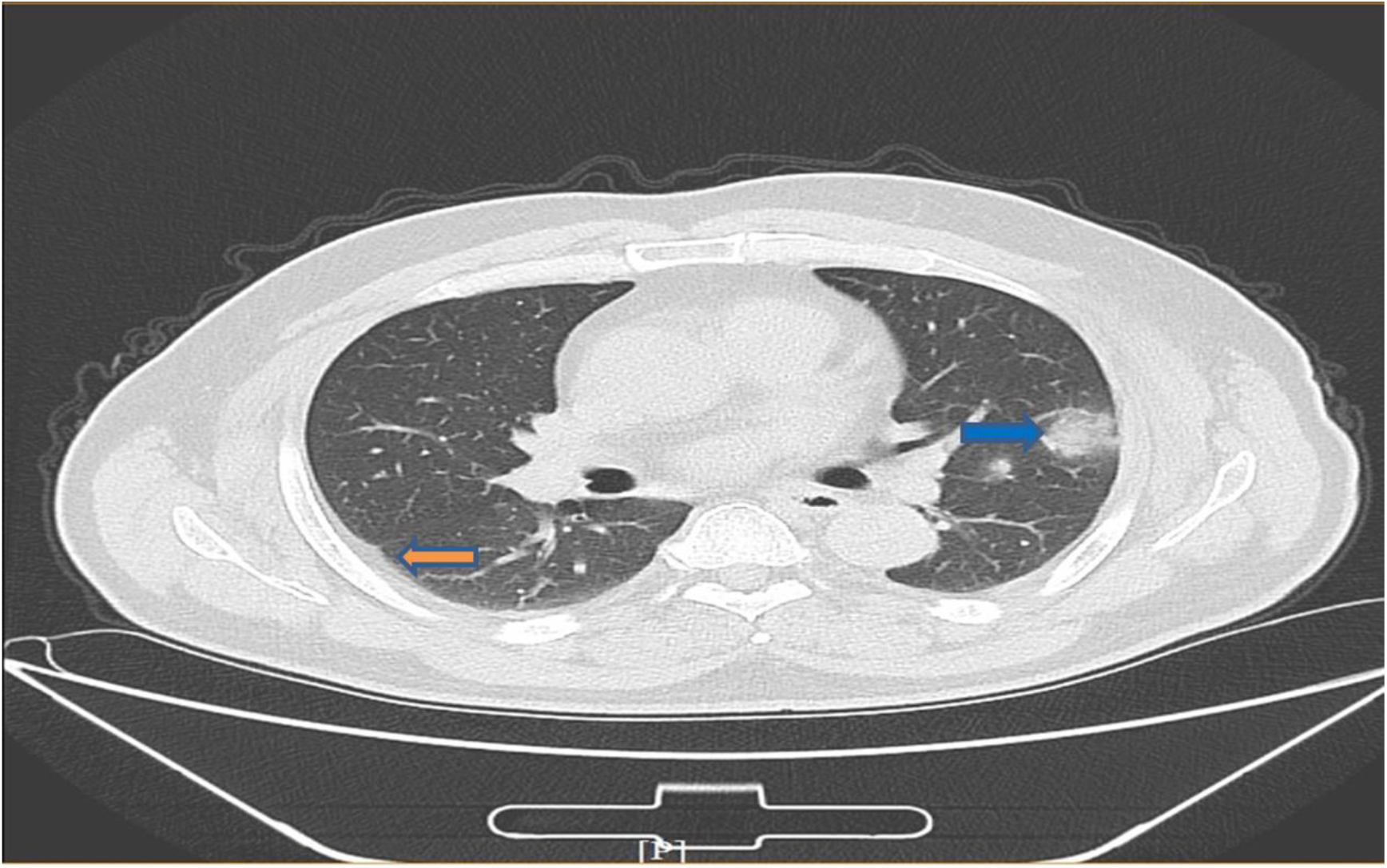
An example of COVID-19 pneumonia features. The blue arrow points to ground-glass opacity, and the yellow arrow points to the pleural indentation sign.

**Fig 3.**
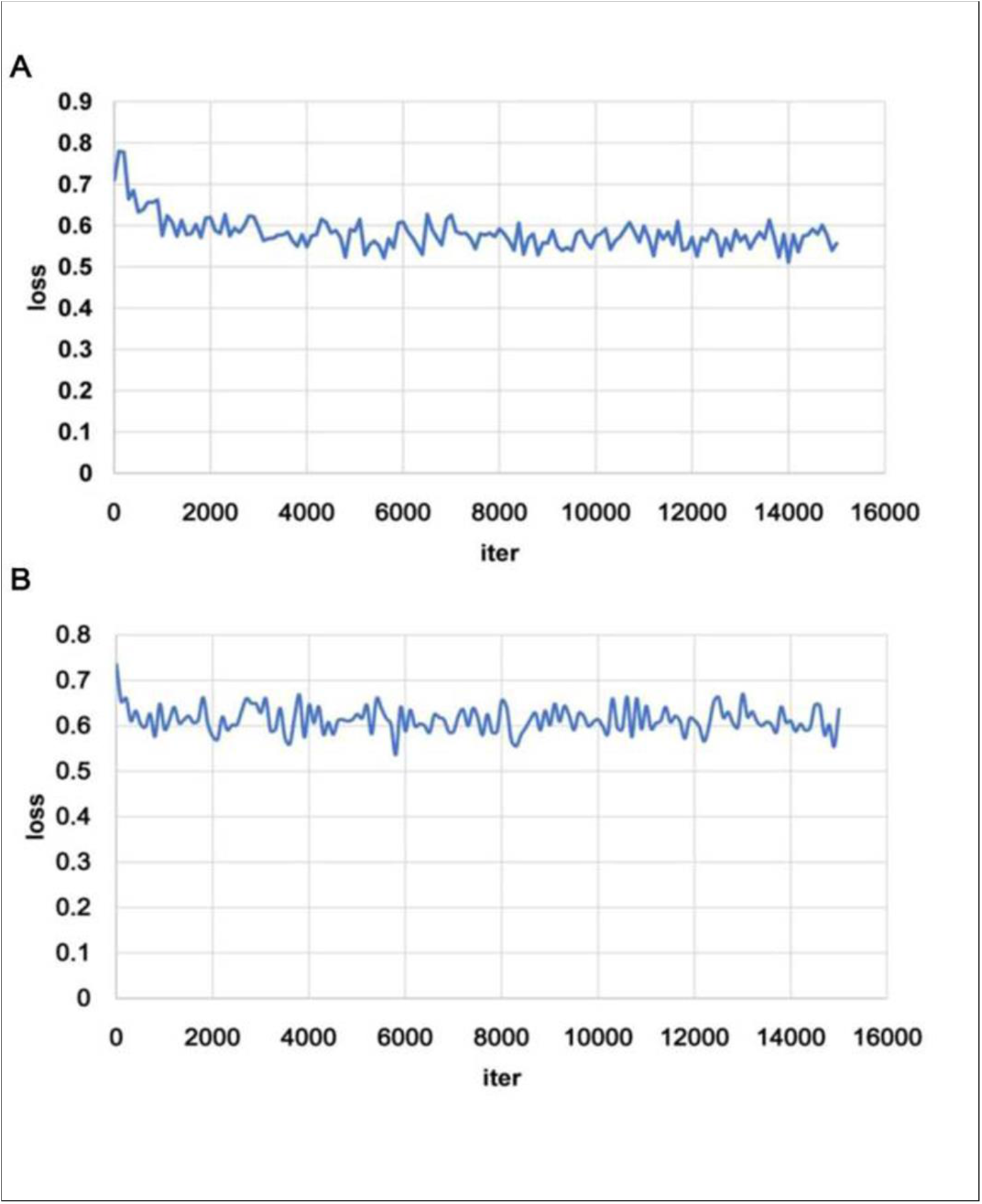
Training loss curves of the models on internal **(A**) and external (**B**) validation. The loss curve tends to be stable after descending, indicating that the training process converges

### Deep learning performance

The deep learning algorithm yielded an AUC of 0.93 (95% CI, 0.90 to 0.96) on the internal validation and 0.81 (95% CI, 0.71 to 0.84) on the external validation based on the certain CT images (**Fig 4)**. Using the maximized Youden index threshold probability, the sensitivity was 0.88 and 0.83, specificity 0.87 and 0.67, the accuracy 89.5% and 79.3%, the negative prediction values 0.95 and 0.90, the Youden indexes 0.75 and 0.48, and the F1 scores were 0.77 and 0.63 for the internal and external datasets, respectively (**Table 2**). The kappa values were 0.69 and 0.48 for internal and external validation in certain CT images, indicating that prediction of COVID-19 from the CNN model is a highly consistent with pathogenic testing results. We also performed an external validation based on each patient’s multiple images. The accuracy was 82.5%, the sensitivity 0.75, the specificity 0.86, the PPV 0.69, the NPV 0.89, and the kappa value was 0.59.

**Table 2.** **Deep learning Algorithm Performance**

| Performance Metric | Internal | External |
| --- | --- | --- |
| AUC (95%CI) | 0.93(0.86 to 0.94) | 0.81(0.71 to 0.84) |
| Accuracy, % | 89.5 | 79.3 |
| Sensitivity | 0.88 | 0.83 |
| Specificity | 0.87 | 0.67 |
| PPV | 0.71 | 0.55 |
| NPV | 0.95 | 0.90 |
| Kappa* | 0.69 | 0.48 |
| Yoden index | 0.75 | 0.50 |
| F1 score‡ | 0.77 | 0.63 |

\* Measures the agreement between the CNN model prediction and the clinical diagnosis. ‡Measures the accuracy of the CNN model.

**Fig 4.**
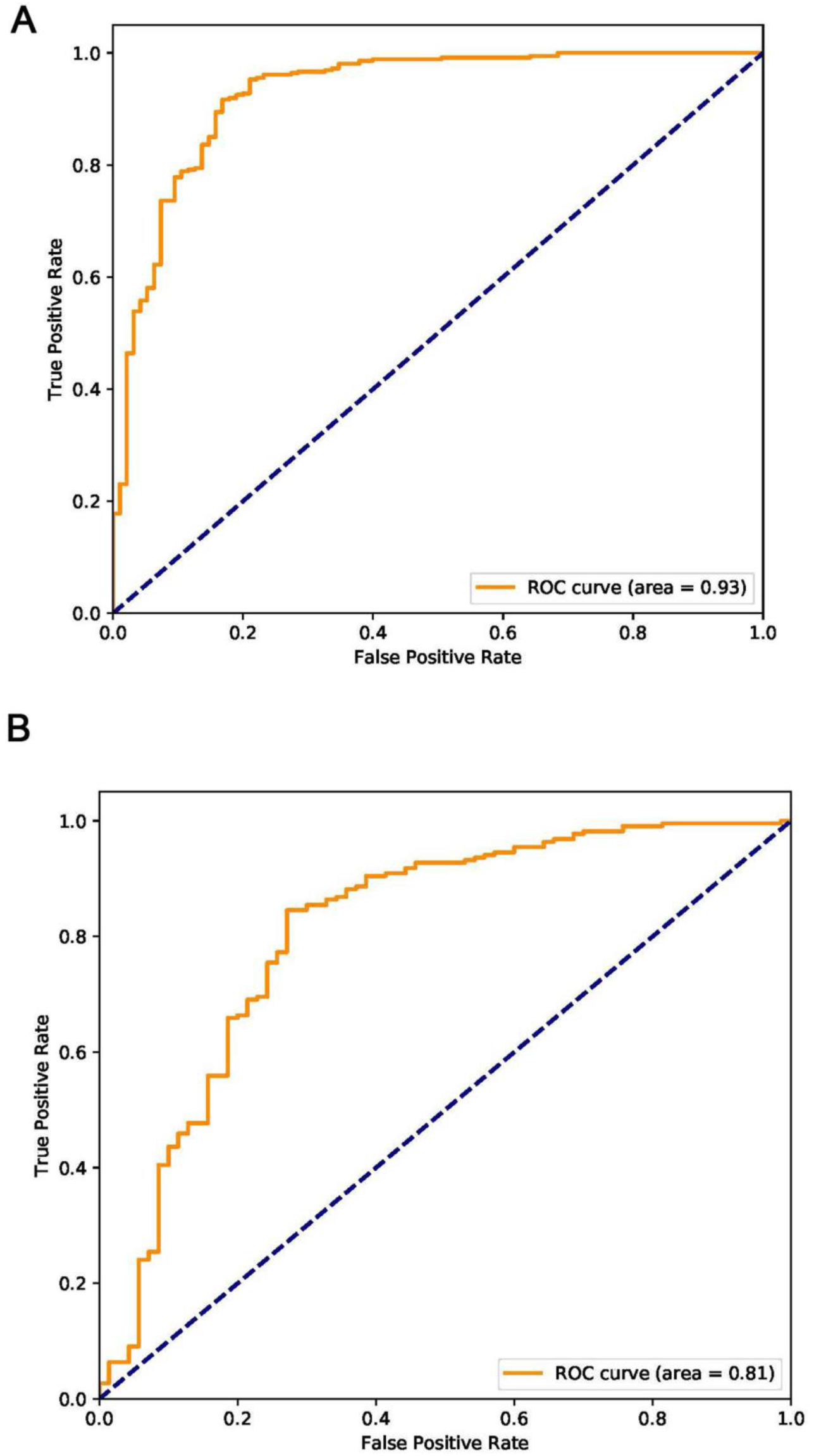
Receiver operating characteristic plots for COVID-19 identification for the deep learning (Inception) algorithm. (A) Internal Validation. (B) External Validation.

### Comparison of AI with radiologist prediction

At the same time, we asked two skilled radiologists to assess the 745 images for a prediction. Radiologist 1 achieved the accuracy of 55.8% with sensitivity of 0.71 and specificity of 0.51, and Radiologist 2 achieved a similar accuracy of 55.4% with sensitivity of 0.73 and specificity of 0.50 (**Table 3**). These results indicates that it is difficult for radiologists to make prediction of COVID-19 with eye recognition, further showing the advantage of the algorithm we developed.

**Table 3.** **Performance metrics for the CNN model versus skilled radiologists**.

| Performance Metric | Internal | External (Based on ROI) | External (Based on patients) | R1 | R2 |
| --- | --- | --- | --- | --- | --- |
| Accuracy, % | 89.5 | 79.3 | 82.5 | 55.8 | 55.4 |
| Sensitivity | 0.88 | 0.83 | 0.75 | 0.71 | 0.73 |
| Specificity | 0.87 | 0.67 | 0.86 | 0.51 | 0.5 |
| PPV | 0.71 | 0.55 | 0.69 | 0.29 | 0.29 |
| NPV | 0.95 | 0.90 | 0.89 | 0.86 | 0.86 |
| F1 score | 0.77 | 0.63 | 0.72 | 0.41 | 0.42 |
| Kappa | 0.69 | 0.48 | 0.59 | 0.15 | 0.15 |
| Yoden index | 0.75 | 0.50 | 0.61 | 0.22 | 0.23 |

### Prediction of COVID-19 on CT images from pathogenic negative patients

Because high false negative results were frequently reported from nucleic acid testing, we aimed to test whether the algorithm could detect COVID-19 when the pathogenic test came negative. To achieve this goal, we enrolled additional 15 COVID-19 cases, in which initial two nucleic acid tests came negative and for the third test they became positive. These CT results were taken on the same day of the nucleic acid tests **(Fig 5**). Interestingly, we found that, 46 out of the 54 images when nucleic acid test results were negative were predicted as COVID-19 positive by the algorithm, with the accuracy of 85.2%. These results indicate that the algorithm has high value serving as a screening method for COVID-19.

**Fig 5.**
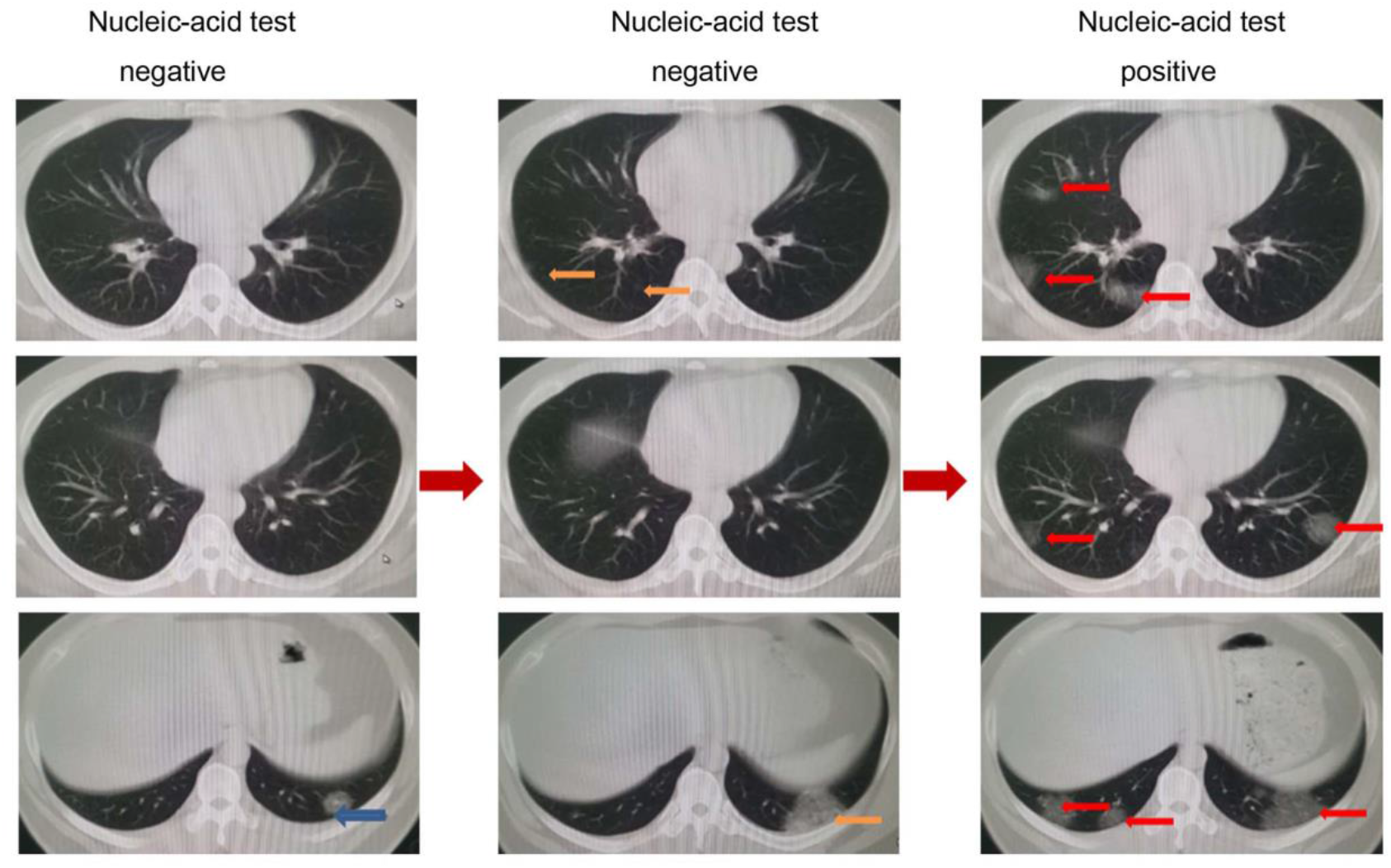
Representative images from a COVID-19 patient with two negatively reported nucleic acid tests at earlier stages and one final positively reported test at a later stage. On the left, only one inflammatory lesion (blue arrow) can be seen near diaphragm. In the middle, lesions (yellow arrows) were found in two levels of images. On the right, the images were taken on the ninth day after admission. The inflammation continued to progress, extending to both lungs (red arrows), and the nucleic acid test became positive.

## Discussion

Timely diagnosis and triaging of PUIs are crucial for the control of emerging infectious diseases such as the current COVID-19. Due to the limitation of nucleic acid -based laboratory testing, there is an urgent need to look for fast alternative methods that can be used by front-line health care personals for quickly and accurately diagnosing the disease. In the present study, we have developed an AI program by analyzing representative CT images using a deep learning method. This is a retrospective, multicohort, diagnostic study using our modified Inception migration neuro network, which has achieved an overall 89.5% accuracy. Moreover, the high performance of the deep learning model we developed in this study was tested using external samples with 79.3% accuracy. More importantly, as a screening method, our model achieved a relative high sensitivity, 0.88 and 0.83 on internal and external certain CT images datasets, respectively. Furthermore, the model achieved a better performance on a certain people, the accuracy up to 82.5%. Of note, our model was used to distinguish between COVID-19 and other typical viral pneumonia, both of which have quite similar radiologic characteristics. During current COVID-19 global pandemics, the CNN model can potentially serve as a powerful tool for COVID-19 screening.

It is important to note that our model aims to distinguish between COVID-19 and other typical viral pneumonia, both of which have similar radiologic characteristics. We compared the performance of our model with that of two skilled radiologists, and our model has shown much higher accuracy and sensitivity. These findings have demonstrated the proof-of-principle that deep learning can extract CT image features of COVID-19 for diagnostic purposes. Using the supercomputer system, the time for each case takes only about 10 seconds, and it can be performed remotely via a shared public platform. Therefore, further developing this system can significantly shorten the diagnosis time for disease control. Our study represents the first study to apply artificial intelligence technologies to CT images for effectively screening for COVID-19.

The gold standard for COVID-19 diagnosis has been nucleic acid based detection for the existence of specific sequences of the SARS-COV-2 gene. While we still value the importance of nucleic acid detection in the diagnosis of SARS-COV-2 infection, we must also note that the high number of false negatives due to several factors such as methodological disadvantages, disease stages, and methods for specimen collection might delay diagnosis and disease control. Recent data have suggested that the accuracy of nucleic acid testing is only about 30-50% [6,7,8]. Using CT imaging feature extraction, we are able to achieve above 89.5% accuracy, significantly outplaying nucleic acid testing. More interestingly, testing CT images from COVID-19 patients when initial pathogenic testing came negative, our model has achieved the accuracy of 85.2% for correctly predicting COVID-19. According to a study authored by Xia L et al, 75% patients with negative RT-PCR results had positive CT findings [17]. The study recommended that chest CT as a primary tool for the current COVID-19 detection in epidemic areas.

Deep learning methods have been used to solve data-rich biology and medicine. A large number of labeled data is required for training [18]. Although we are satisfied with the initial results, we believe that with more CT images included in the training, we will achieve higher accuracy. Therefore, further optimizing and testing this system is warranted. To achieve this, we have generated a webpage that licensed healthcare personnel can access to upload CT images for testing and validation. The webpage information is as following: https://ai.nscc-tj.cn/thai/deploy/public/pneumonia_ct.

There are some limitations to our study. Although DL has been used to represent and learn predictable relationships in many diverse forms of data, and it holds promise for applications in precision medicine, many factors such as low signal to noise and complex data integration have challenged the DL efficacy [19]. CT images represent a difficult classification task due to the relatively large number of variable objects, specifically the imaged areas outside the lungs that are irrelevant to the diagnosis of pneumonia [12]. In addition, the training data set is relatively small. The performance of this system is expected to increase when the training volume is increased. It should also be noted that, the features of the CT images we analyzed were from patients with severe lung lesions at later stages of disease development. Although we have enrolled 15 cases of COVID patients for assessing the value of the algorithm for early diagnosis, larger numbers of database to associate this with the disease progress and all pathologic stages of COVID-19 is necessary to optimize the diagnostic system.

In future, we intend to link hierarchical features of CT images to features of other factors such as genetic, epidemiological and clinical information for multi-modeling analysis for an enhanced diagnosis. The artificial intelligence system developed in our study should significantly contribute to COVID-19 disease control by reducing the number of PUIs for timely quarantine and treatment.

## Ethics Committee Approval and Patient Consent

This study complies with the Institutional Review Board of each participating institutes. Informed consent was exempted by the IRB because of the retrospective nature of this study.

## Data Availability

All data referred to in the manuscript is available.

https://ai.nscc-tj.cn/thai/deploy/public/pneumonia_ct

## Abbreviations

COVID-19: Corona Virus Disease
CT: Computed Tomography
SARS-COV-2: severe acute respiratory syndrome coronavirus 2Convolutional
CNN: Neural Network
ROI: region of interest
M-Inception: modified Inception
AUC: Area Under Curve
PPV: Positive predictive value
NPV: Negative predictive value
CV: computer vision

## Reference

1. Zhou F, Yu T, Du R, Fan G, Liu Y, Liu Z, et al. Clinical course and risk factors for mortality of adult inpatients with COVID-19 in Wuhan, China: a retrospective cohort study. Lancet. Mar. 2020.

2. Velavan T, Meyer C. The COVID-19 epidemic. Trop Med Int Health, Mar 2020.

3. Wang D, Hu B, Hu C, Zhu F, Liu X, Zhang J, et al. Clinical Characteristics of 138 Hospitalized Patients With 2019 Novel Coronavirus-Infected Pneumonia in Wuhan, China. JAMA. 2020

4. Chen N, Zhou M, Dong X, Qu J, Gong F, Han Y, et al. Epidemiological and clinical characteristics of 99 cases of 2019 novel coronavirus pneumonia in Wuhan, China: a descriptive study. Lancet. 2020.

5. Li Q, Guan X, Wu P, Wang X, Zhou L, Tong Y, et al. Early Transmission Dynamics in Wuhan, China, of Novel Coronavirus-Infected Pneumonia. N Engl J Med. Mar. 2020.

6. Huang C, Wang Y, Li X, Ren L, Zhao J, Hu Y, et al. Clinical features of patients infected with 2019 novel coronavirus in Wuhan, China. Lancet. 2020.

7. Corman VM, Landt O, Kaiser M, Molenkamp R, Meijer A, Chu DK, et al. Detection of 2019 novel coronavirus (2019-nCoV) by real-time RT-PCR. Euro surveillance vol. 25, Jan. 2020.

8. Chu DKW, Pan Y, Cheng SMS, Hui KPY, Krishnan P, Liu Y, et al. Molecular Diagnosis of a Novel Coronavirus (2019-nCoV) Causing an Outbreak of Pneumonia. Clinical chemistry. 2020.

9. Zhang N, Wang L, Deng X, Liang R, Su M, He C, et al. Recent advances inthe detection of respiratory virus infection in humans. J Med Virol. 2020.

10. Chung M, Bernheim A, Mei X, Zhang N, Huang M, Zeng X, et al. CT Imaging Features of 2019 Novel Coronavirus (2019-nCoV). Radiology. Apr. 2020.

11. Gomez P, Semmler M, Schutzenberger A, Bohr C, Dollinger M. Low-light image enhancement of high-speed endoscopic videos using a convolutional neural network. Med Biol Eng Comput. 2019; 57(7): 1451–63.

12. Choe J, Lee SM, Do KH, Lee G, Lee JG, Lee SM, et al. Deep Learning-based Image Conversion of CT Reconstruction Kernels Improves Radiomics Reproducibility for Pulmonary Nodules or Masses. Radiology. 2019; 292(2): 365–73.

13. Kermany DS, Goldbaum M, Cai W, Valentim CCS, Liang H, Baxter SL, et al.Identifying Medical Diagnoses and Treatable Diseases by Image-Based Deep Learning. Cell 2018; 172(5): 1122–31.

14. Negassi M, Suarez-Ibarrola R, Hein S, Miernik A, Reiterer A. Application of artificial neural networks for automated analysis of cystoscopic images: a review of the current status and future prospects. World J Urol. 2020.

15. Wang P, Xiao X, Glissen Brown JR, Berzin TM, Tu M, Xiong F, et al.Development and validation of a deep-learning algorithm for the detection of polyps during colonoscopy. Nat Biomed Eng. 2018; 2(10): 741–8.

16. Koo HJ, Lim S, Choe J, Choi SH, Sung H, Do KH. Radiographic and CT Features of Viral Pneumonia. Radiographics. 2018; 38(3): 719–39.

17. Ai T, Yang Z, Hou H, Zhan C, Chen C, Lv W, et al. Correlation of Chest CT and RT-PCR Testing in Coronavirus Disease 2019 (COVID-19) in China: A Report of 1014 Cases [published online ahead of print, 2020 Feb 26]. Radiology. 2020; p.200642.

18. Ching T, Himmelstein DS, Beaulieu-Jones BK, Kalinin AA, Do BT, Way GP, et al. Opportunities and obstacles for deep learning in biology and medicine. J R Soc Interface. 2018;15(141):20170387. doi:10.1098/rsif.2017.0387

19. Grapov D, Fahrmann J, Wanichthanarak K, Khoomrung S. Rise of Deep Learning for Genomic, Proteomic, and Metabolomic Data Integration in Precision Medicine. OMICS. 2018;22(10):630–636.

